# The effect of variation of individual infectiousness on SARS-CoV-2 transmission in households

**DOI:** 10.1101/2022.08.30.22279377

**Authors:** Tim K. Tsang, Xiaotong Huang, Can Wang, Sijie Chen, Bingyi Yang, Simon Cauchemez, Benjamin J. Cowling

## Abstract

Quantifying variation of individual infectiousness is critical to inform disease control. Previous studies reported substantial heterogeneity in transmission of many infectious diseases (including SARS-CoV-2). However, those results are difficult to interpret since the number of contacts is rarely considered in such approaches. Here, we analyze data from 17 SARS-CoV-2 household transmission studies conducted in periods dominated by ancestral strains, in which the number of contacts was known. By fitting individual-based household transmission models to these data, accounting for number of contacts and baseline transmission probabilities, the pooled estimate suggests that the 20% most infectious cases have 3.1-fold (95% confidence interval: 2.2-4.2 fold) higher infectiousness than average cases, which is consistent with the observed heterogeneity in viral shedding. Household data can inform the estimation of transmission heterogeneity, which is important for epidemic management.

**One Sentence Summary:** In this study, variation of individual infectiousness is quantified. Potential sources of such variation, particularly heterogeneity of viral shedding is discussed.

## INTRODUCTION

Characterizing transmission is critical to control the spread of an emerging infectious disease. The reproductive number is the widely adopted measure of infectiousness. However, it only measures the average number of secondary cases infected by an infected person, not the heterogeneity in the number of transmissions. Variation of individual infectiousness is particularly highlighted by superspreading events (SSEs), in which a minority of cases are responsible for a majority of transmission events. Such phenomena, illustrated by the “80/20 rule” (i.e., 20% of cases responsible for 80% transmission (*1, 2*)), have been observed in emerging infectious disease outbreaks (*3*), including severe acute respiratory syndrome (SARS) (*4*), middle east respiratory syndrome (MERS) (*5, 6*) and most recently the COVID-19 pandemic (*1, 2, 7-9*). In these outbreaks, the proportion of cases attributed to 80% transmission (*p_80_*), and the dispersion parameter (*k*) were estimated by fitting the negative binomial distribution to the number of secondary cases (*3*) as a measure of transmission heterogeneity.

However, the number of contacts per index cases is often not reported in SSE studies, and hence not incorporated in the analyses. In addition, SSE studies usually analyze clusters from different settings, in which the baseline transmission risk and density of exposure could be different (*10*). Finally, studies of transmission heterogeneity that focus on SSEs described in the literature may suffer from publication bias, with larger clusters having higher probability of being observed and reported (*8*). Therefore, the observed heterogeneity in the number of secondary cases could be a result of large number of contacts in SSE settings, or confounding from these factors (*11, 12*), instead of variation in individual infectiousness.

Households are one of the most important settings for SARS-CoV-2 transmission, with 4-fold to 10-fold higher transmission risk than other places (*10*). Hence, household transmission studies provide an ideal setting to quantify variations in individual infectiousness. In a household transmission study, an index case is identified, and their household contacts are followed-up for one to two weeks, during which there is high transmission potential (*13*). Therefore, the number of contacts is known while transmission risks and reporting biases can be controlled. We aim to characterize the variation of individual infectiousness by analyzing data from household transmission studies.

## RESULTS

### Estimating the variation of individual infectiousness from household studies

We conducted a systematic review to gather information on the number of secondary cases with the number of household contacts for each household, in the form of number of households with *X* cases among households of size *Y*. In total, we identified 17 studies, comprising 13098 index cases and 31359 household contacts (Fig. S1, Table S1) (*14–30*). Most studies covered the cases from January to November 2020, which was dominated by ancestral strains, except for Layan et al. (from December 2020 to April 2021) and Hsu et al. (from January 2020 to February 2021), which covered both ancestral strains and the alpha variant.

We then developed a statistical model to quantify the degree and the impact of variation of infectiousness of cases on transmission dynamics. The individual-based household transmission model describes the probability of infection of household contacts as depending on the time since infection in other infected persons in the household, so that infections from outside the household (community infections), or infections via other household contacts rather than the index case (tertiary infections) are allowed (*31–33*). We extend this model by adding a random effect (*δ_i_*) on the individual infectiousness of cases. Here, the relative infectiousness of case *i* compared with case *j* is exp(*δ_i_*)/exp(*δ_j_*). The parameter for variation in individual infectiousness (hereafter denoted as infectiousness variation, *σ_var_*) is the standard deviation (SD) of the random effect characterizing individual infectiousness, so that *δ_i_* follows Normal distribution with mean equal to 0 and SD equal to *σ_var_*.

We separately fit the models to 14 studies with more than 150 contacts (*14–27*) (Fig. 1, Table S2). For 12 studies out of 14, models with infectiousness variation perform substantially better (range of ΔDIC: 5.8-268) (Figure 2, Table S2). From these 12 studies, the estimated infectiousness variation (*σ_var_*) ranged from 1.03 to 2.83. This suggests that, the 20% most infectious cases are 2.4-fold to 10-fold more infectious than the average case. Based on the two largest studies with 6782 and 3727 households (*14, 15*), the estimated infectiousness variation is 1.48 (95% credible interval (CrI): 1.29, 1.7) and 1.41 (95% CrI: 1.19, 1.72), suggesting that, among all cases, the 20% most infectious are 3.5-fold (95% CrI: 3.0-4.2 fold) and 3.3-fold (95% CrI: 2.7-4.3 fold) more infectious than the average case. The estimated daily probability of infection from outside the household and estimated person-to-person transmission probability within households are ranged from 0.0003 to 0.017, and from 0.06 to 0.51 respectively. The estimates of parameters for the relationship between number of contacts and transmission (larger value indicates stronger inverse association) are ranged from 0.43 to 0.92, except for the study by Layan et al (*18*) where it is equal to 0.2. For all studies, the predicted final size distribution is consistent with the observed data and the model fit is judged adequate (Table S3-S4).

**Fig. 1.**
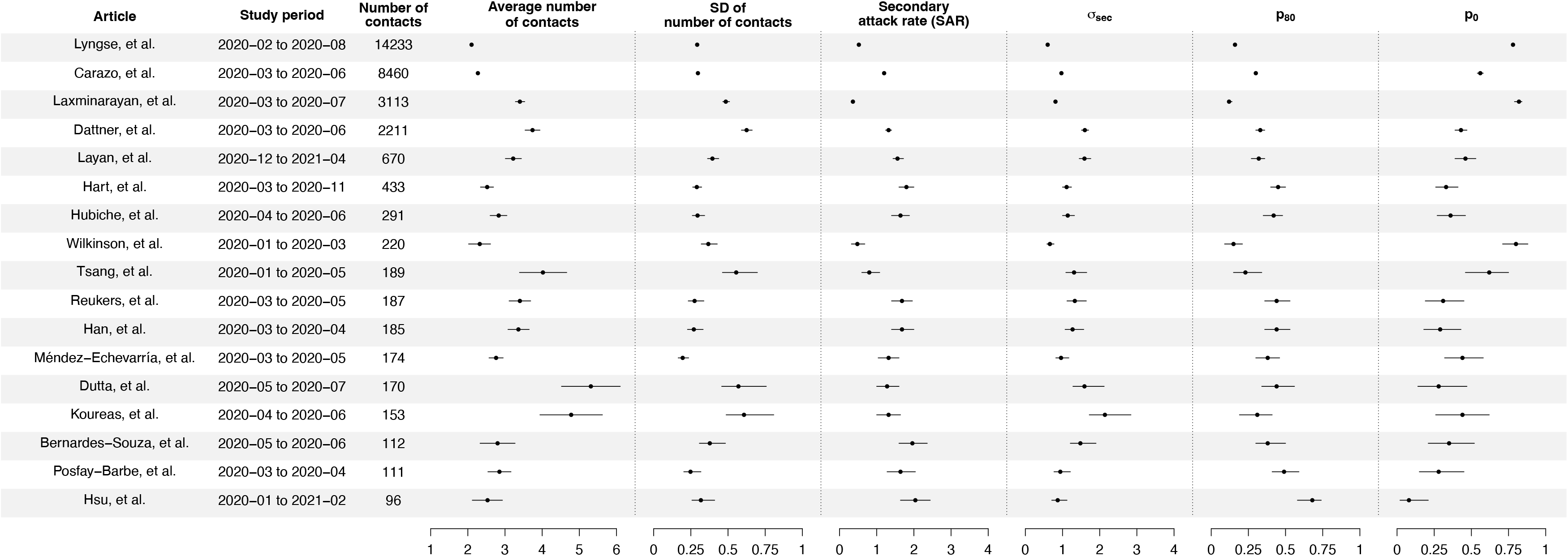
Summary of statistics for 17 identified studies. Figure shows the average number of contacts and SD of number of contact, SD of number of secondary cases per index cases (*σ_sec_*), proportion of households in which no contacts are infected (*p_80_*), proportion of cases attributed to 80% transmission (*p_80_*) and secondary attack rate (SAR) for 17 identified studies.

**Fig. 2.**
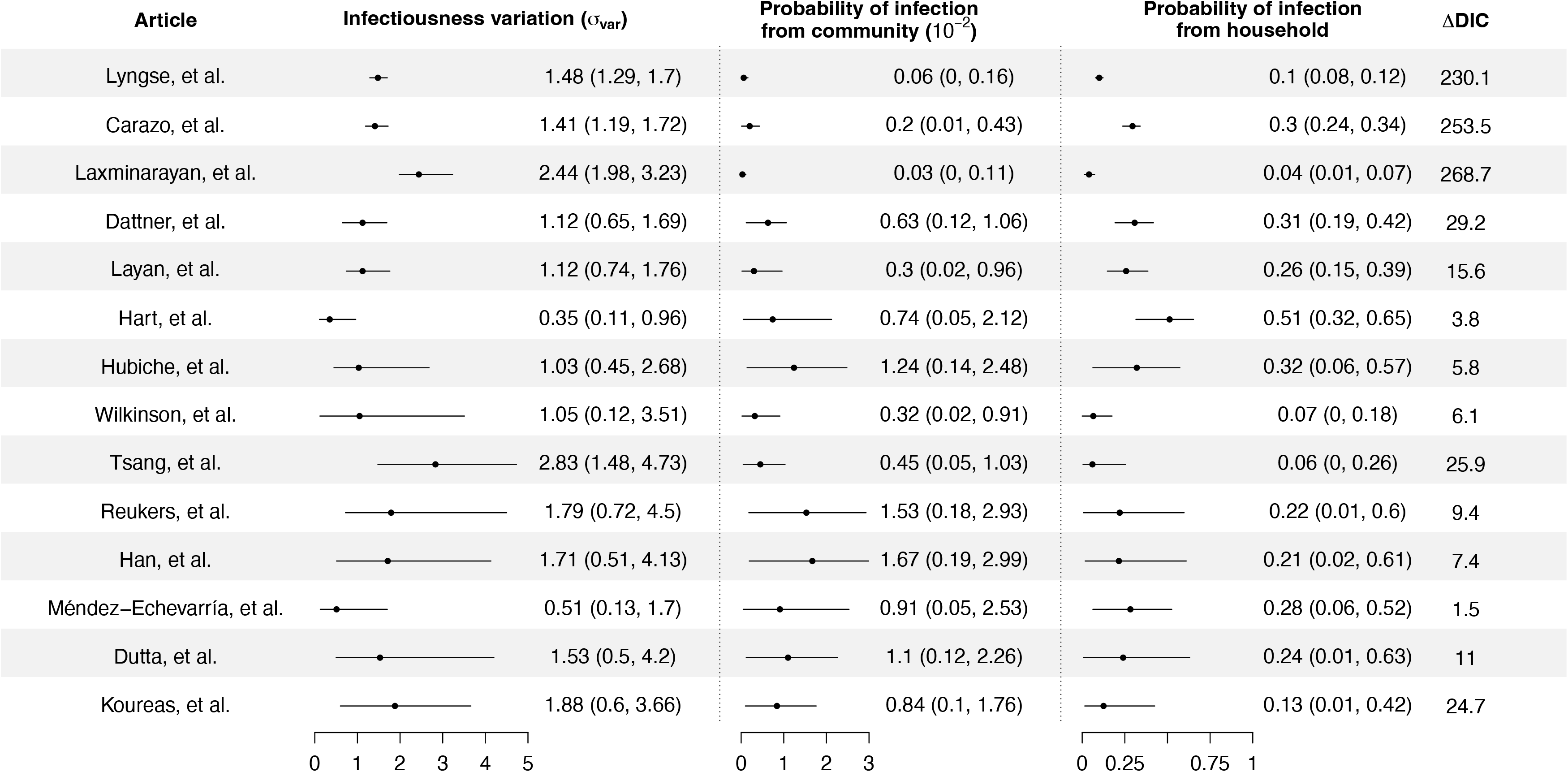
Modeling results of household transmission dynamics and infectiousness variation. Figure shows the estimates of infectiousness variation (*σ_var_*), the estimated probability of infection from community and estimated probability of infection from households, and the reduction in DIC compared with the model without infectiousness variation. Models are fitted separately to 14 household transmission studies.

We conduct random effects meta-analyses on estimates of individual infectiousness from 14 identified studies. The pooled estimate of infectiousness variation is 1.33 (95% confidence interval (CI): 0.95, 1.70), suggesting that the 20% most infectious cases are 3.1-fold (95% CI: 2.2-4.2 fold) more infectious than the average case (Figure 4). Based on this fitted distribution, we estimate that 5.9% (95% CI: 1.4%, 11.1%) and 14.9% (95% CI: 7.2%, 20.7%) of cases could be at least 8-fold and 4-fold more infectious than average cases, respectively.

**Fig. 3.**
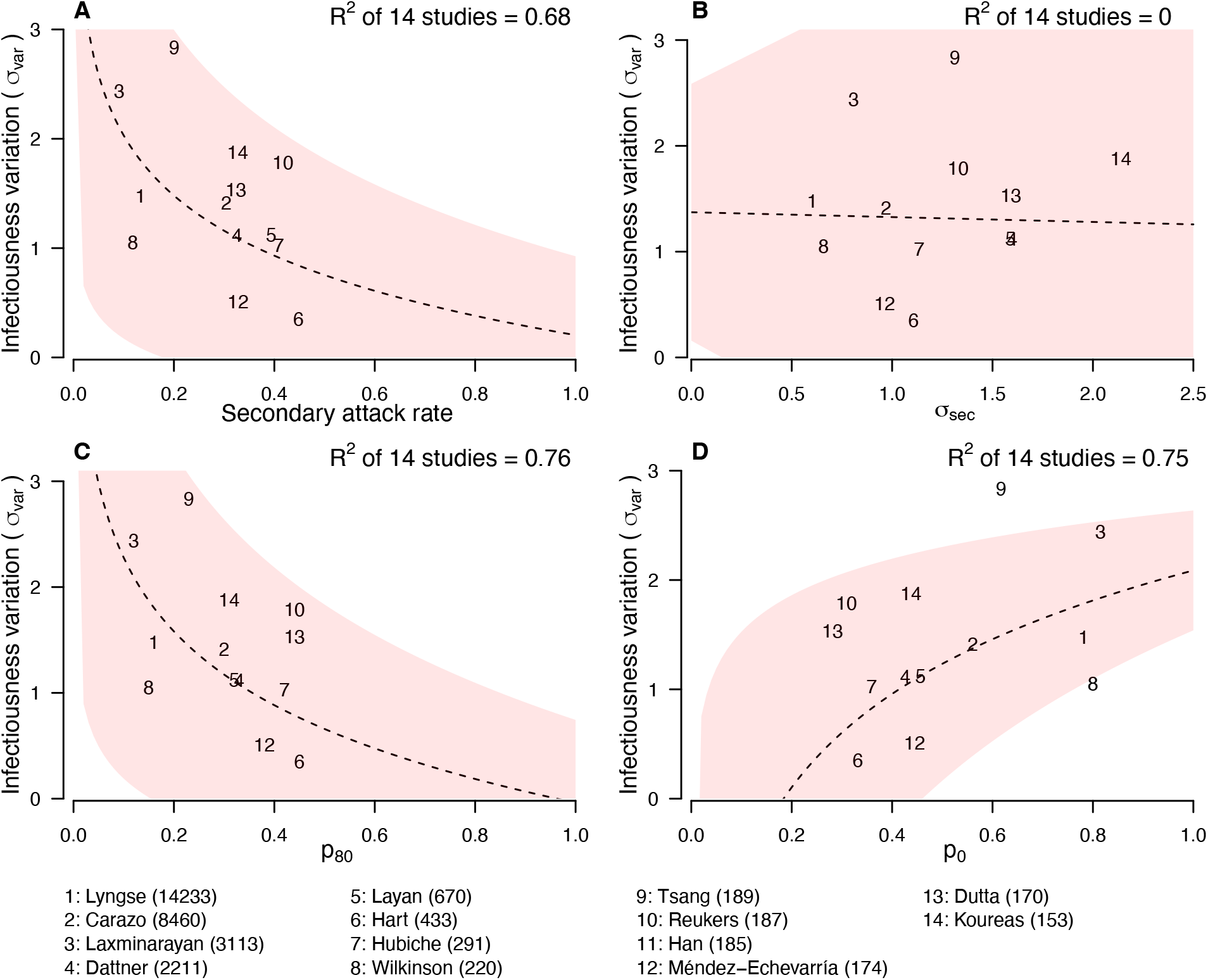
Relationship between infectiousness variation and statistic. In each panel, numbers represent the observed corresponding relationship for the identified studies. Panel A, B, C and D show the relationship between infectiousness variation (*σ_var_*) and secondary attack rate (SAR), proportion of households in which no contacts are infected (*p_0_*), proportion of cases attributed to 80% transmission (*p_80_*) and SD of number of secondary cases per index cases (*σ_sec_*). In the bottom, numbers in bracket indicate the number of household contacts in corresponding studies.

**Fig. 4.**
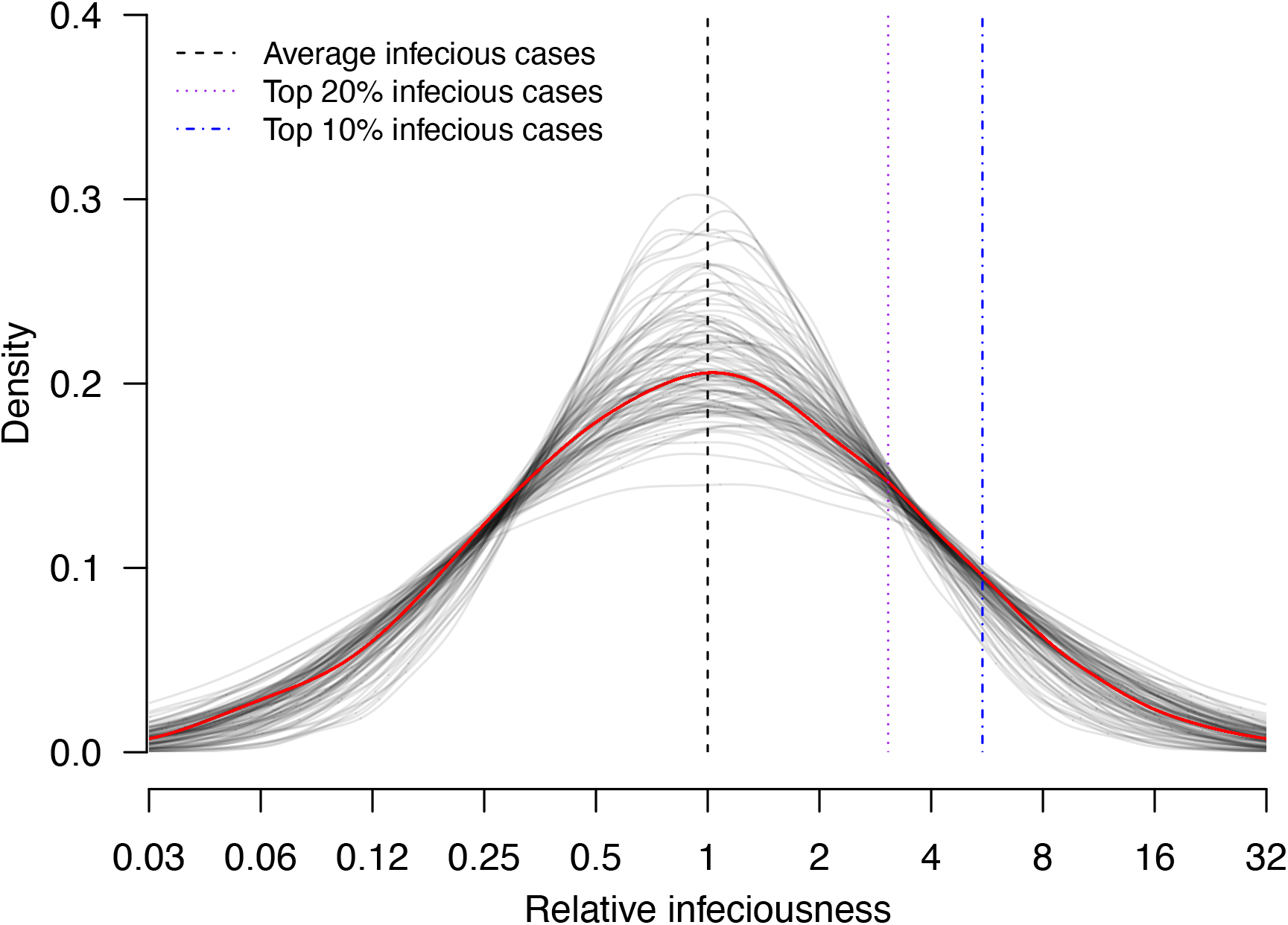
Estimate distribution of relative infectiousness based on the pooled estimate. Red line indicates the estimated distribution and the gray lines indicate the associated uncertainty. Black dashed line indicates average infectiousness (relative infectiousness equal to 1), while the purple and blue dashed lines indicate top 20% and 10% infectiousness respectively.

### Proxy measures of infectiousness variation

While such a modeling approach generates valuable information on infectiousness variation from household studies, directly applying this approach may be challenging due to the complexity in modelling and estimation. There is therefore a need to determine the relationship between characteristic of household transmissions and infectiousness variation, to design and validate a proxy of infectiousness variation, such as the widely used *p_80_*. We consider the following candidate: 1) *p_80_*, defined as the proportion of index cases that had >80% of their household contacts becoming cases in the study, 2) the proportion of households in which no contacts are infected (*p_0_*), 3) the secondary attack rate (SAR, the proportion of infected contacts), and 4) the standard deviation (SD) of the distribution of number of secondary cases (*σ_sec_*) (Fig. 2; Table S5).

In meta-regression, we find the infectiousness variation is associated with *p*_80_, *p*_0_ and SAR. We estimate that doubling *p*_80_and SAR are associated with 0.70 (95% CI: 0.30, 1.09) and 0.55 (95% CI: 0.21, 0.89) unit decrease in infectiousness variation, with R-squared equal to 76% and 68% respectively. We also estimate that doubling *p*_0_is associated with 0.85 (95% CI: 0.33, 1.38) unit increase in infectious variation (R^2^ = 75%). In addition, we find that higher infectiousness variation is associated with only using PCR to ascertain secondary cases. Regarding other statistics, we find that *σ_sec_* is positively associated with mean and SD of number of contacts. Regarding other factors, we find that higher infectiousness variation is associated with only using PCR to confirm secondary cases. Other than these associations, we find no association between these statistics and implementation of lockdown, ascertainment method of index and secondary cases, and the circulating virus of SARS-CoV-2 in the study period.

## DISCUSSION

In this study, we characterize the impact of variation of individual infectiousness on heterogeneity of transmission of COVID-19 in households. We demonstrate that it can be estimated from household data using a modeling approach. The pooled estimate of infectiousness variation from 14 studies suggests that the 20% most infectious cases have 3.1-fold (95% CI: 2.2-4.2-fold) higher infectiousness compared with average index cases. This implies there is substantial variation in individual infectiousness of cases in households. This variation could be attributed to both biological factors and host behaviours.

Regarding host behaviours, multiple contact patterns, particularly by age, could contribute to the variations in infectiousness of cases. For example, mother-child contacts are usually more intense than father-child contacts, and adult cases are more capable of self-isolation within a household compared with children. Furthermore, contact pattern studies suggested that school-age children and young adults tended to mix with people of the same age (*34, 35*). Biological factors may also contribute to such variations. For example, the SAR for index cases with fever and cough are 1.4-fold and 1.3-fold higher than index cases without fever and cough respectively (*36*). Viral shedding is used as a proxy measure of infectiousness, and consistently it also has substantial variations (*37*). Regarding the magnitude of viral shedding, studies report high variations of temporal viral shedding patterns among individuals (*38–40*). In addition, the duration of viral shedding can be highly heterogeneous, with pooled estimates of mean durations ranging from 11.1 days to 30.3 days, and almost all reviews report high heterogeneity of estimates identified from literatures (*37, 41–50*). Such heterogeneities still exist in subgroup analyses by age and severity (*37, 41-45, 50*). Also, the infectious period, proxied by duration of replicant competent virus isolation, is also heterogeneous (*50*). One review also suggest that heterogeneity in viral shedding is an intrinsic virological factor facilitating higher dispersion parameter for SARS-CoV-2 if we compare it with the corresponding patterns in SARS-CoV-1 and pandemic influenza A(H1N1)pdm09 (*51*).

The observed variation in individual infectiousness is consistent with past analyses of the dispersion parameter in a negative binomial distribution fitted to number of secondary cases per index case of COVID-19 (*2, 52*). However, in these studies, the number of contacts are not considered. Therefore, the observed variations may not apply directly to households with limited number of contacts. Here, we explore the relationship between infectiousness variations and household characteristics, and the commonly adopted measure of heterogeneity, proportion of cases attributed to 80% transmission (*p_80_*). We find that infectiousness variation is strongly associated with *p_80_*, similar to previous analysis about superspreading, suggesting that it could be a measure of infectiousness variation in households.

Infectiousness variation is also correlated with the secondary attack rate (SAR), and the proportion of households in which no contacts are infected (*p_0_*). When secondary attack rate is higher, it is expected that more contacts are infected and therefore the observed number of secondary cases would be less heterogeneous. The strong association between *p_0_* and infectiousness variation suggests that the observed variations may also be attributed to some cases that are less infectious than average, which is consistent with Hong Kong data (*53*). In contrast, the SD of the distribution of number of secondary cases (*σ_sec_*) is only weakly correlated with infectiousness variation. This is because *σ_sec_* is highly correlated with the mean and SD of number of contacts, suggesting that it may depend on distribution of number of contacts, and hence may not be comparable among studies. We find higher infectiousness variation is associated with only using PCR to confirm secondary cases. One potential reason is that using other methods may lower the sensitivity of detecting infection, resulting in lower estimates of SAR and hence higher estimates of infectiousness variation. Further investigations are needed to explore roles of other potential factors affecting infectiousness variation, such as contact frequency among different regions (*35*).

An important limitation of our study is that we do not have individual-level data. Therefore, we are unable to determine the impact of demographic factors like age and sex on infectiousness variation. Also, we cannot disentangle the host behaviors from biological factors. Previous analysis suggested no evidence of the impact of age on infectiousness of cases (*2, 9*). In addition, we could not include factors affecting susceptibility to infection. Our estimates of infectiousness variation should be interpreted in light of these limitations: they capture heterogeneity in infectiousness due to demographic, host and biological factors. If we were able to account for these factors thanks to the analysis of more detailed datasets, our estimates of infectiousness variations would likely be lower. However, in one study that included susceptibility component in the estimation of individual infectiousness, substantial heterogeneity remained with 20% of cases estimated to contribute to 80% of transmission (*22*). Second, the recruitment methods among studies are different. This may affect the comparability of the results, although all index cases are laboratory-confirmed in all studies (*32*). Finally, most of our identified studies were conducted in the period of circulation of ancestral strains, and therefore the identified infectiousness variation may not be directly applicable to other variants.

In conclusion, we developed a modeling approach to estimate variation in individual infectiousness from household data. Result indicates that there is substantial variation in individual infectiousness, which is important for epidemic management.

## MATERIALS AND METHODS

### Study design

The aim of this study was to develop a statistical model to quantify the variation of individual infectiousness in households, based on publicly available information. An index case was defined as the first detected case in a household, while secondary cases were defined as the identified infected household contacts of the index case. We conducted a systematic review to collect household studies with at least 30 households, reporting the number of secondary cases with number of household contacts for each household for COVID-19, in the form of number of households with *X* cases among households of size *Y*. For each study, we also extracted the study period, the coverage of tests of household contacts, the case ascertainment methods, the circulating virus of SARS-CoV-2, and the public health and social measures in the study period. This information was used as an input of modeling analyses in this study. Details of systematic review could be found in Materials and Methods Section 1.

### Estimation of variation of individual infectiousness in households

To determine if there are variations of individual infectiousness of cases, we use an individual-based household transmission model (*31–33*). The model describes the probability of infection of household contacts as depending on the time since infection in other infected people in the household, while infections from outside the household (community infections), or infections via other household contacts rather than the index case (tertiary infections) are allowed. We extend the model by adding a random effect parameter (*δ_i_*) on the individual infectiousness of each case. Here, *δ_i_* follows a normal distribution with mean 0 and standard deviation *σ_var_*, which quantify the variation of individual infectiousness (hereafter denoted as infectiousness variation). The relative infectiousness of case *i* compared with case *j* is exp(*δ_i_*)/exp(*δ_j_*).

Since the data were extracted from publication, the infection time of cases was unavailable. Also, the individual infectiousness parameters *δ_i_* for cases were augmented variables. Therefore, we conducted our inference under a Bayesian framework with using data-augmentation Markov chain Monte Carlo (MCMC) algorithm to joint estimate the model parameters, the missing infection time and augmented variables with using metropolis-hasting algorithm. We separately fitted this model to identified studies. We assessed the model adequacy by comparing the observed and expected number of infections in households by household size. We compared the model with or without the random effect for variation of individual infectiousness by using Deviance Information Criterion (DIC) (*54*). Smaller DIC indicated a better model fit. DIC difference >5 was considered as substantial improvement (*55*). Details of the model and inference could be found in Materials and Methods Section 2 and 3.

### Statistical analysis

We conducted random effects meta-analyses on identified studies to obtain pooled estimates of individual infectiousness and *p_80_*, using the inverse variance method and restricted maximum likelihood estimator for heterogeneity (*56–59*). Cochran *Q* test and the *I^2^* statistic were used to identify and quantify heterogeneity among included studies (*60, 61*). An *I^2^* value more than 75% indicates high heterogeneity (*61*). We conducted a meta regression analysis to explore the association between these statistics, and further including the following factors: the mean and SD of number of household contacts, implementation of lockdown, ascertainment method of index and secondary cases, only ancestral strains are circulating in study period, and all household contacts were tested.

## Data Availability

The data and computer code (in R languages) for conducting the data analysis can be downloaded from https://github.com/timktsang/covid19_transmission_heterogeneity

## Acknowledgments

The authors thank Maylis Layan for helpful discussion and Jiayi Zhou for technical assistance.

## Author Contributions

Tim K. Tsang: Conceptualization, Methodology, Software, Data curation, Writing-Original draft preparation, Visualization, Validation

Xiaotong Huang: Data curation, Investigation, Visualization

Can Wang: Data curation, Investigation

Sijie Chen: Investigation, Visualization

Bingyi Yang: Conceptualization, Writing-Reviewing and Editing

Simon Cauchemez: Conceptualization, Methodology, Supervision, Writing-Reviewing and Editing

Benjamin J. Cowling: Conceptualization, Methodology, Supervision, Writing-Reviewing and Editing

## Supplementary Materials

### 1 Systematic review

We conducted a systematic review following the Preferred Reporting Items for Systematic Review and Meta-analysis (PRISMA) statement (*62*). A standardized search was done in PubMed, Embase and Web of Science, using the search term “(“COVID-19” OR “SARS-CoV-2”) AND (“household” OR “family”) AND (“transmission”)”. The search was done on 22 June, 2022, with no language restrictions. Additional relevant articles from the reference sections were also reviewed.

Two authors (XH and CW) independently screened the titles and extracted data from the included studies, with disagreement resolved by consensus together with a third author (TKT). Studies identified from different databases were de-duplicated.

### 2 Household transmission model

Our aim was to explore the role of variation of individual infectiousness on transmission, with accounting for the variation and number of contacts, and the transmission probability. Therefore, we extended the household transmission model to account for variation of individual infectiousness of index cases (*31*). We added a random effect on infectiousness of cases (*33*). In the model, the hazard of infection of individual *j* at time *t* from an infected household member *i,* with infection time *t_i_* in household *k*, was

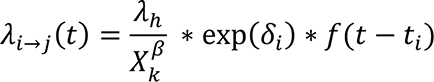

where *λ_h_* was the baseline hazard, *δ_i_* was the relative individual infectiousness of cases, which followed a normal distribution with mean 0 and standard deviation *σ_var_*.

*X_k_* was the number of household contacts. *β* was the parameter describing the relationship between number of household contacts and transmission rate. It ranged from 0 to 1, with 0 indicating that the transmission rate is independent of number of household contacts while 1 indicates that the transmission rate is inversely proportional to number of household contacts (i.e. dilution effect of the contact time per contact which is lower when the number of household contacts is higher). *f(.)* was the infectiousness profile since infection generated from the assumed incubation period (mean equal to 5 days) and infectious period (mean equal to 13) (*40, 63, 64*).

The daily hazard of infection from outside the household is assumed to be *λ_c_*(*t*) = *λ_c_*. Hence, the total hazard infection of individual j on day t was

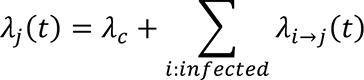

Based on the transmission model, the probability that an individual *i* was infected with infection time *t_i_*_0_ was

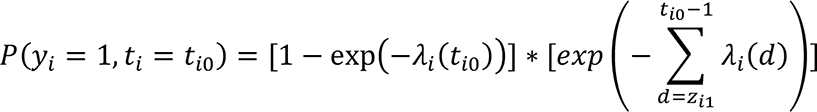

where *z_i_*_1_ was the start of the follow-up day of individual *i*. Denote the total follow-up time for an individual *i* as *z_i_*_2_, then the probability that an individual *i* did not get infected within the follow-up period is

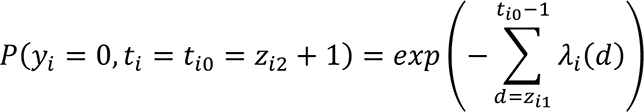

Hence the log-likelihood function L was

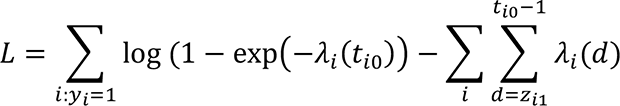

Index cases did not contribute to the likelihood and hence the summation was only on household contacts.

### 3 Inference, model comparison and validation

The infection time for cases were unavailable in the data extracted from publication. Also, the individual infectiousness parameter *δ_i_* was an augmented variable. Therefore, we conducted our inference under a Bayesian framework with using data-augmentation Markov chain Monte Carlo (MCMC) algorithm to joint estimate the model parameters, the missing infection time and the relative individual infectiousness with using metropolis-hasting algorithm. For the parameter that only takes positive value, we used a vague Uniform(0,10) prior. For the parameter for relationship between number of household contacts and transmission, we used Uniform(0,1). The algorithm runs for 50000 iterations after a burn-in of 50000 iterations. Converge was visually assessed. One run takes about 3 hours on our typical desktop computer. We fitted the model to all studies, but only 14 of them could converge and provide robust estimates of individual infectiousness.

We assessed the model adequacy by comparing the observed and expected number of infections in households by household size. We simulated 10000 datasets with a structure identical to that of the observed data (in terms of number of household contacts), with parameters randomly drawn from the posterior distribution of model parameter.

We compared the model with or without the random effect for variation of individual infectiousness by using Deviance Information Criterion (DIC) (*54*). Smaller DIC indicated a better model fit. DIC difference >5 was considered as substantial improvement (*55*). Since the likelihood of observed data could not be directly evaluated for a given model (*65*), we used an importance sampling approach to estimate the likelihood for the observed data and evaluate the DIC (*66, 67*). For each household, we simulated 2000 datasets, with parameters randomly drawn from the posterior distribution. Then we compared the observed data and simulated data. The contribution to the likelihood of a household was equal to the proportion of simulated data with infection status that exactly matched the observed data, for all household members. To avoid the problem of 0-valued likelihood, we used the approach developed by Cauchemez et al (*66*), and assumed the sensitivity and specificity for diagnosing a case were both 99.99%.

---

**Fig. S1.**
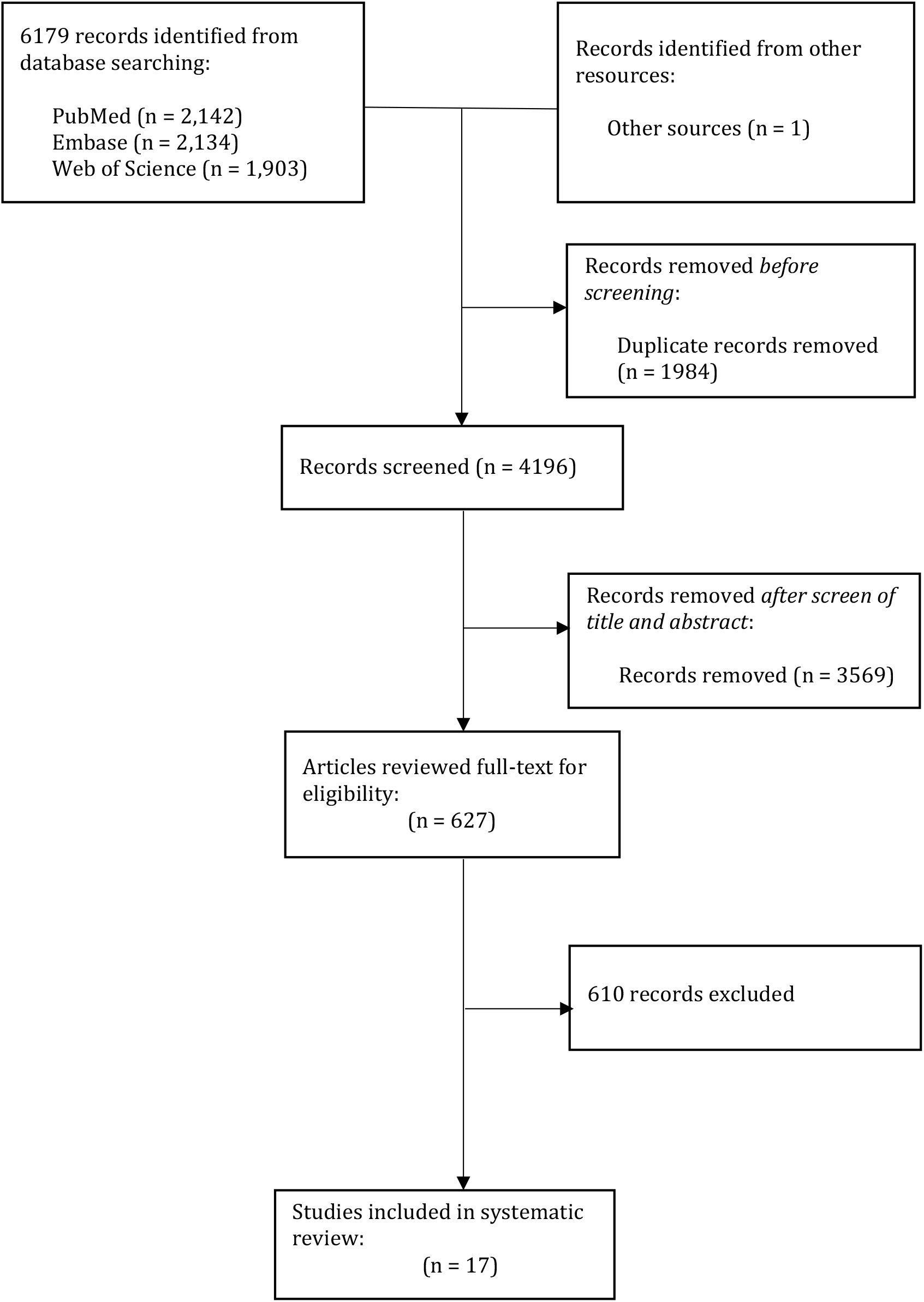
Process of systematic review

**Table S1.**
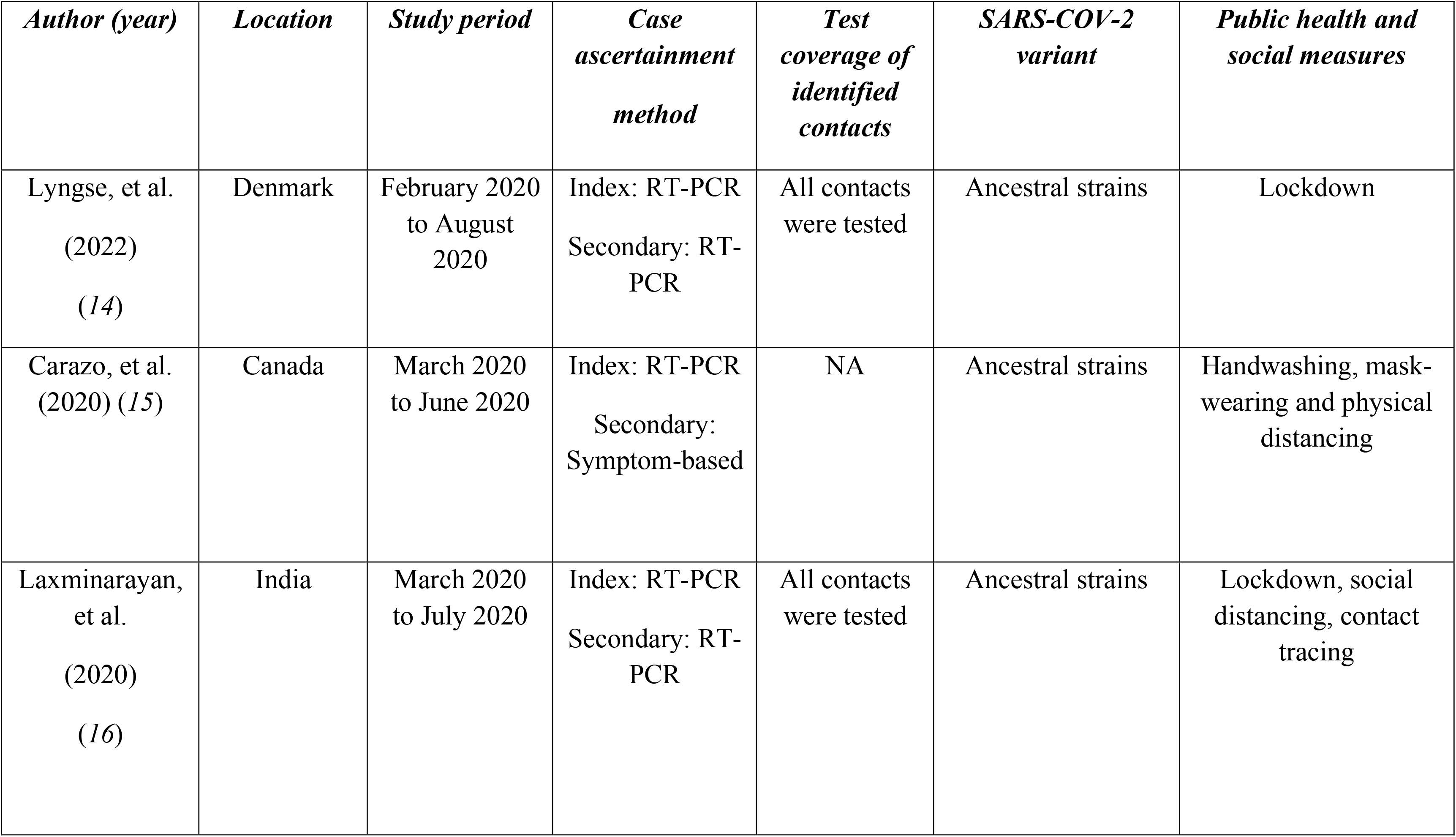
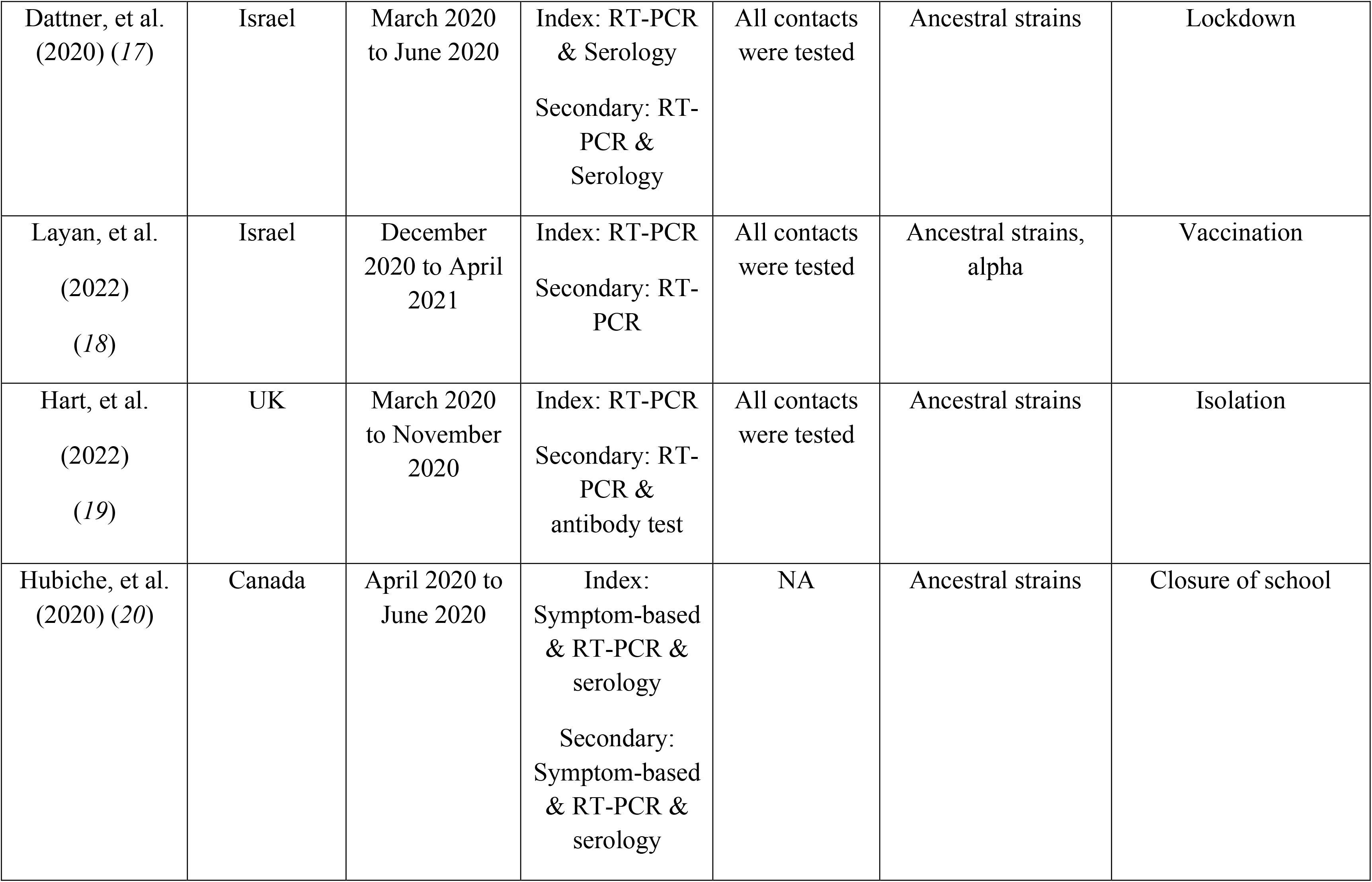
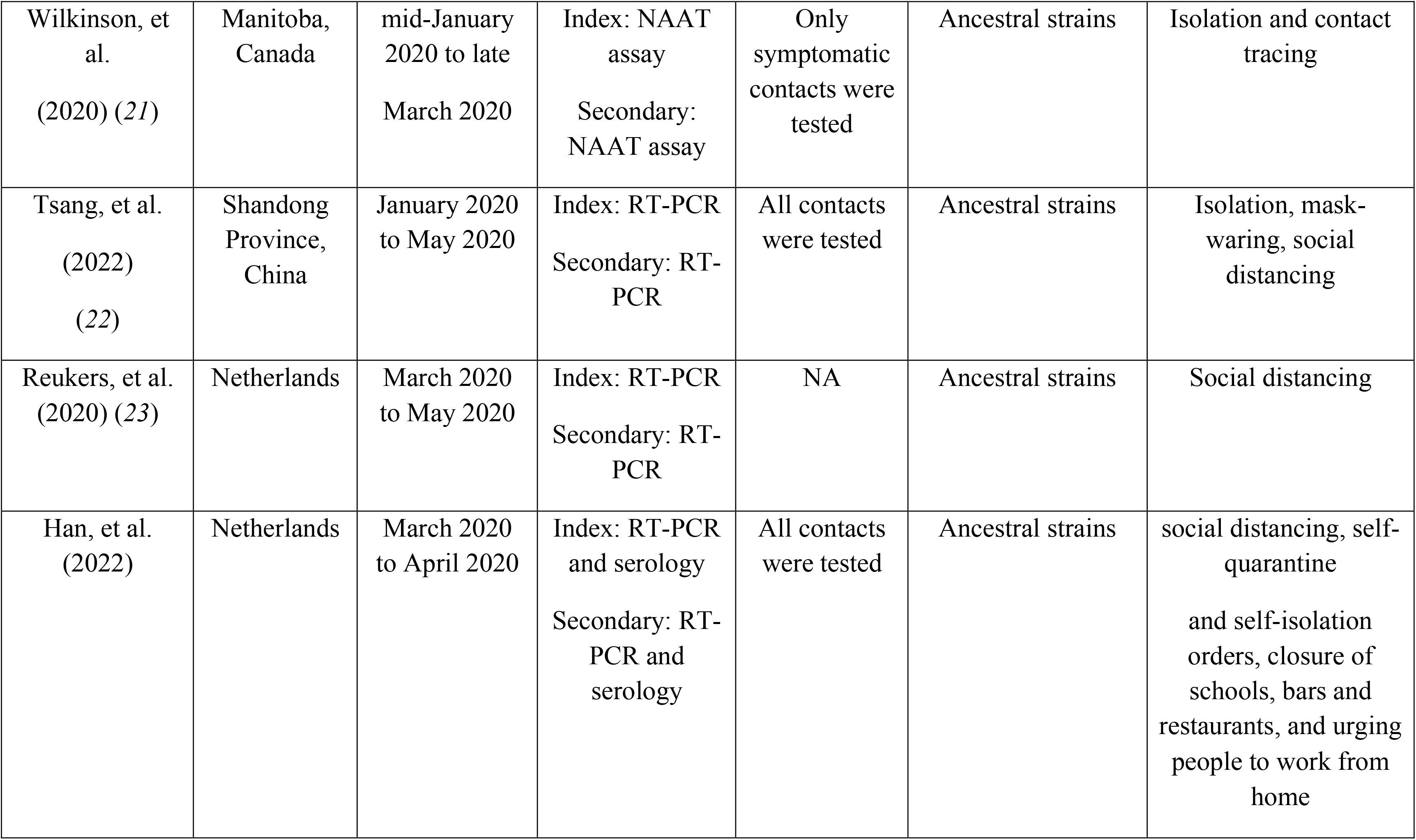
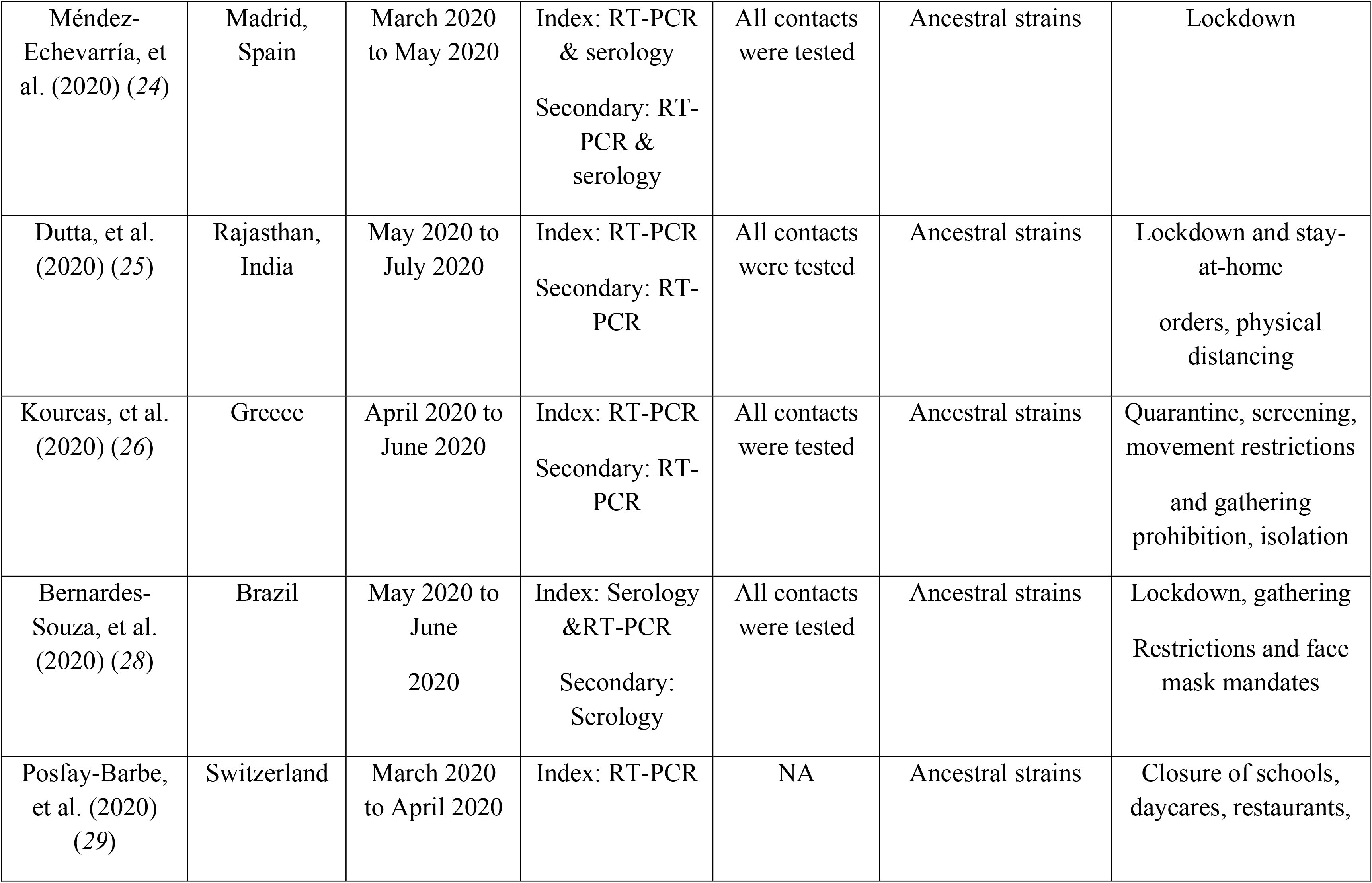
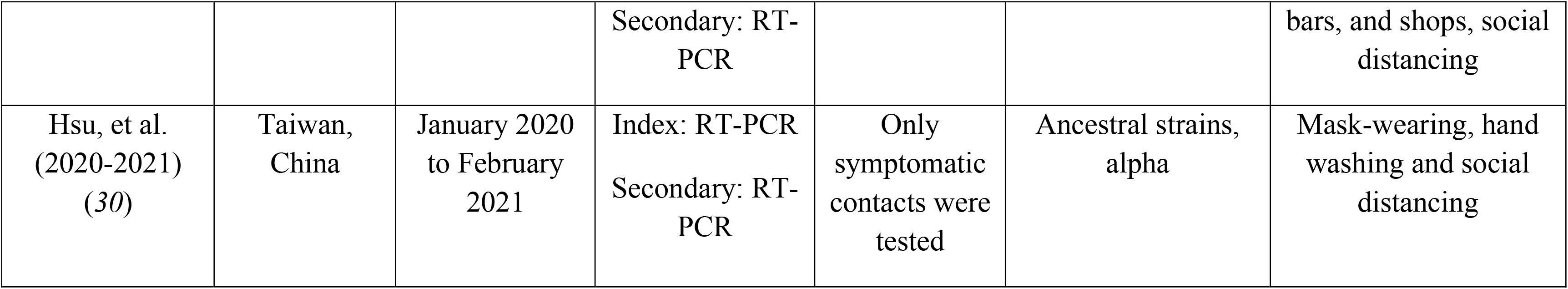
Summary of characteristic of identified studies.

**Table S2.** Summary of model estimates

| Article | Estimates of infectiousness variation | Estimates of probability of infection from community ($10^{-2}$) | Estimates of probability of infection from household | Relationship between transmission and number of contacts ($\beta$ ) |
| --- | --- | --- | --- | --- |
| Lyngse, et al. | 1.48 (1.29, 1.7) | 0.06 (0, 0.16) | 0.1 (0.08, 0.12) | 0.72 (0.59, 0.89) |
| Carazo, et al. | 1.41 (1.19, 1.72) | 0.2 (0.01, 0.43) | 0.3 (0.24, 0.34) | 0.75 (0.62, 0.93) |
| Laxminarayan, et al. | 2.44 (1.98, 3.23) | 0.03 (0, 0.11) | 0.04 (0.01, 0.07) | 0.92 (0.69, 1) |
| Dattner, et al. | 1.12 (0.65, 1.69) | 0.63 (0.12, 1.06) | 0.31 (0.19, 0.42) | 0.65 (0.45, 0.91) |
| Layan, et al. | 1.12 (0.74, 1.76) | 0.3 (0.02, 0.96) | 0.26 (0.15, 0.39) | 0.2 (0.01, 0.6) |
| Hart, et al. | 0.35 (0.11, 0.96) | 0.74 (0.05, 2.12) | 0.51 (0.32, 0.65) | 0.72 (0.32, 0.98) |
| Hubiche, et al. | 1.03 (0.45, 2.68) | 1.24 (0.14, 2.48) | 0.32 (0.06, 0.57) | 0.67 (0.13, 0.98) |
| Wilkinson, et al. | 1.05 (0.12, 3.51) | 0.32 (0.02, 0.91) | 0.07 (0, 0.18) | 0.43 (0.02, 0.97) |
| Tsang, et al. | 2.83 (1.48, 4.73) | 0.45 (0.05, 1.03) | 0.06 (0, 0.26) | 0.66 (0.06, 0.99) |
| Reukers, et al. | 1.79 (0.72, 4.5) | 1.53 (0.18, 2.93) | 0.22 (0.01, 0.6) | 0.67 (0.07, 0.98) |
| Han, et al. | 1.71 (0.51, 4.13) | 1.67 (0.19, 2.99) | 0.21 (0.02, 0.61) | 0.69 (0.07, 0.99) |
| Méndez-Echevarría, et al. | 0.51 (0.13, 1.7) | 0.91 (0.05, 2.53) | 0.28 (0.06, 0.52) | 0.57 (0.04, 0.98) |
| Dutta, et al. | 1.53 (0.5, 4.2) | 1.1 (0.12, 2.26) | 0.24 (0.01, 0.63) | 0.78 (0.15, 0.99) |
| Koureas, et al. | 1.88 (0.6, 3.66) | 0.84 (0.1, 1.76) | 0.13 (0.01, 0.42) | 0.44 (0.02, 0.96) |

**Table S3.** Model adequacy check for Lyngse et al. Each element of the table has the format “observed frequency – expected (posterior mean) frequency (95% Credible interval).

|  | **Number of infected household contacts** |  |  |  |  |  |
| --- | --- | --- | --- | --- | --- | --- |
| **Number of household contacts** | **0** | **1** | **2** | **3** | **4** | **5** |
| **1** | 2366 - 2377<br>(2319, 2433) | 569 - 558 (502, 616) | NA | NA | NA | NA |
| **2** | 1117 - 1105<br>(1070, 1138) | 227 - 227 (198, 259) | 77 - 88 (69, 109) | NA | NA | NA |
| **3** | 1135 - 1118<br>(1077, 1156) | 214 - 235 (203, 267) | 89 - 86 (67, 106) | 41 - 41 (27, 56) | NA | NA |
| **4** | 521 - 528 (501, 555) | 119 - 115 (94, 137) | 40 - 42 (30, 56) | 25 - 19 (11, 30) | 11 - 10 (4, 18) | NA |
| **5** | 161 - 167 (152, 181) | 42 - 38 (27, 50) | 14 - 14 (7, 22) | 7 - 7 (2, 12) | 7 - 3 (0, 8) | 0 - 2 (0, 5) |

**Table S4.**
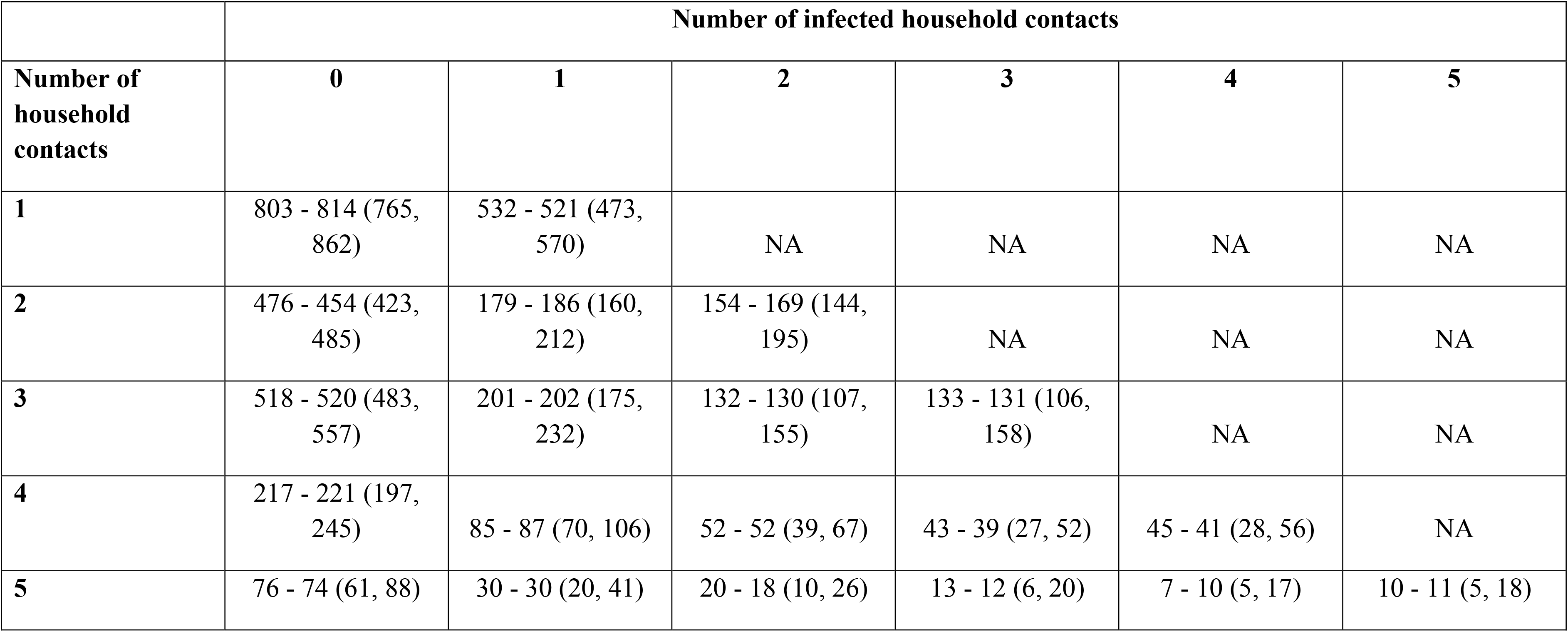
Model adequacy check for Lyngse et al. Each element of the table has the format “observed frequency – expected (posterior mean) frequency (95% Credible interval).

|  | **Number of infected household contacts** |  |  |  |  |  |
| --- | --- | --- | --- | --- | --- | --- |
| **Number of household contacts** | **0** | **1** | **2** | **3** | **4** | **5** |
| **1** | 803 - 814 (765, 862) | 532 - 521 (473, 570) | NA | NA | NA | NA |
| **2** | 476 - 454 (423, 485) | 179 - 186 (160, 212) | 154 - 169 (144, 195) | NA | NA | NA |
| **3** | 518 - 520 (483, 557) | 201 - 202 (175, 232) | 132 - 130 (107, 155) | 133 - 131 (106, 158) | NA | NA |
| **4** | 217 - 221 (197, 245) | 85 - 87 (70, 106) | 52 - 52 (39, 67) | 43 - 39 (27, 52) | 45 - 41 (28, 56) | NA |
| **5** | 76 - 74 (61, 88) | 30 - 30 (20, 41) | 20 - 18 (10, 26) | 13 - 12 (6, 20) | 7 - 10 (5, 17) | 10 - 11 (5, 18) |

**Table S5.** Summary of characteristic of identified studies. SD: standard deviation.

| Article | Number of households | Number of contacts | Number of secondary cases | Mean number of contacts | SD of number of contact | Secondary attack rate (SAR) | SD of number of secondary cases ($\sigma_{sec}$) | Proportion of household in which no contacts are infected ($p_0$) | Proportion of cases attributed to 80% transmission ($p_{80}$) |
| --- | --- | --- | --- | --- | --- | --- | --- | --- | --- |
| Lyngse, et al. | 6782 | 14233 | 1902 | 2.1 (2.07, 2.13) | 1.17 (1.15, 1.19) | 0.13 (0.13, 0.14) | 0.6 (0.59, 0.61) | 0.78 (0.77, 0.79) | 0.16 (0.15, 0.17) |
| Carazo, et al. | 3727 | 8460 | 2574 | 2.27 (2.23, 2.31) | 1.19 (1.16, 1.21) | 0.3 (0.29, 0.31) | 0.97 (0.95, 0.99) | 0.56 (0.54, 0.58) | 0.3 (0.29, 0.31) |
| Laxminarayan, et al. | 915 | 3113 | 283 | 3.4 (3.28, 3.53) | 1.94 (1.86, 2.04) | 0.09 (0.08, 0.1) | 0.81 (0.77, 0.85) | 0.82 (0.79, 0.84) | 0.12 (0.11, 0.13) |
| Dattner, et al. | 591 | 2211 | 720 | 3.74 (3.54, 3.94) | 2.5 (2.36, 2.65) | 0.33 (0.31, 0.35) | 1.6 (1.51, 1.7) | 0.43 (0.39, 0.47) | 0.33 (0.3, 0.36) |
| Layan, et al. | 208 | 670 | 264 | 3.22 (3.01, 3.44) | 1.58 (1.45, 1.75) | 0.39 (0.36, 0.43) | 1.59 (1.45, 1.76) | 0.46 (0.39, 0.53) | 0.32 (0.27, 0.37) |
| Hart, et al. | 172 | 433 | 194 | 2.52 (2.34, 2.69) | 1.16 (1.05, 1.29) | 0.45 (0.4, 0.5) | 1.11 (1, 1.24) | 0.33 (0.26, 0.41) | 0.45 (0.4, 0.5) |
| Hubiche, et al. | 103 | 291 | 119 | 2.83 (2.6, 3.05) | 1.18 (1.04, 1.37) | 0.41 (0.35, 0.47) | 1.14 (1, 1.32) | 0.36 (0.27, 0.46) | 0.42 (0.35, 0.49) |
| Wilkinson, et al. | 95 | 220 | 26 | 2.32 (2.02, 2.61) | 1.47 (1.28, 1.71) | 0.12 (0.08, 0.17) | 0.66 (0.58, 0.77) | 0.8 (0.71, 0.88) | 0.15 (0.09, 0.21) |
| Tsang, et al. | 47 | 189 | 38 | 4.02 (3.39, 4.66) | 2.22 (1.85, 2.79) | 0.2 (0.15, 0.27) | 1.31 (1.09, 1.65) | 0.62 (0.46, 0.75) | 0.23 (0.15, 0.31) |
| Reukers, et al. | 55 | 187 | 78 | 3.4 (3.11, 3.69) | 1.1 (0.93, 1.35) | 0.42 (0.35, 0.49) | 1.33 (1.12, 1.64) | 0.31 (0.19, 0.45) | 0.44 (0.36, 0.52) |
| Han, et al. | 55 | 185 | 78 | 3.36 (3.08, 3.65) | 1.08 (0.91, 1.33) | 0.42 (0.35, 0.5) | 1.27 (1.07, 1.57) | 0.29 (0.18, 0.43) | 0.44 (0.36, 0.52) |
| Méndez-Echevarría, et al. | 63 | 174 | 57 | 2.76 (2.57, 2.95) | 0.78 (0.66, 0.94) | 0.33 (0.26, 0.4) | 0.96 (0.82, 1.17) | 0.44 (0.32, 0.58) | 0.38 (0.3, 0.46) |
| Dutta, et al. | 32 | 170 | 55 | 5.31 (4.52, 6.1) | 2.28 (1.83, 3.03) | 0.32 (0.25, 0.4) | 1.59 (1.28, 2.12) | 0.28 (0.14, 0.47) | 0.44 (0.34, 0.54) |
| Koureas, et al. | 32 | 153 | 50 | 4.78 (3.94, 5.62) | 2.43 (1.95, 3.23) | 0.33 (0.25, 0.41) | 2.14 (1.72, 2.84) | 0.44 (0.26, 0.62) | 0.31 (0.19, 0.43) |
| Bernardes-Souza, et al. | 40 | 112 | 55 | 2.8 (2.33, 3.27) | 1.51 (1.23, 1.93) | 0.49 (0.4, 0.59) | 1.48 (1.21, 1.9) | 0.35 (0.21, 0.52) | 0.38 (0.3, 0.46) |
| Posfay-Barbe, et al. | 39 | 111 | 46 | 2.85 (2.54, 3.16) | 0.99 (0.81, 1.27) | 0.41 (0.32, 0.51) | 0.94 (0.77, 1.21) | 0.28 (0.15, 0.45) | 0.49 (0.41, 0.57) |
| Hsu, et al. | 38 | 96 | 49 | 2.53 (2.12, 2.93) | 1.27 (1.03, 1.64) | 0.51 (0.41, 0.61) | 0.87 (0.71, 1.12) | 0.08 (0.02, 0.21) | 0.68 (0.58, 0.78) |

**Table S6.** Association between infectiousness variation estimated from household transmission models, and other statistics from 17 household studies, based on meta-regression.

| Statistic | Infectiousness variation ($\sigma_{var}$)* | Standard deviation (SD) of the distribution of number of secondary cases ($\sigma_{sec}$) | Proportion of households in which no contacts are infected ($p_0$) | Proportion of cases attributed to 80% transmission ($p_{80}$) | Secondary attack rate (SAR) |
| --- | --- | --- | --- | --- | --- |
| Pooled estimates | 1.33 (0.95, 1.70) | 1.19 (1.03, 1.35) | 0.39 (0.30, 0.49) | 0.37 (0.30, 0.45) | 0.35 (0.28, 0.44) |
| $I^2$ | 78.4 | 99.2 | 99.5 | 98.8 | 99.1 |
| Factors |  |  |  |  |  |
| Estimate | $\beta$ | $\beta$ | $\exp(\beta)$ | $\exp(\beta)$ | $\exp(\beta)$ |
| Mean number of contacts | 0.31 (-0.16, 0.78) | **0.35 (0.22, 0.48)** | 0.85 (0.63, 1.14) | 1.09 (0.86, 1.38) | 1.09 (0.83, 1.43) |
| SD number of contacts | 0.55 (-0.12, 1.21) | **0.42 (0.15, 0.69)** | 1.04 (0.64, 1.71) | 0.79 (0.54, 1.16) | 0.80 (0.51, 1.25) |
| Implementation of Lockdown | 0.16 (-0.61, 0.93) | -0.06 (-0.40, 0.28) | 1.34 (0.79, 2.28) | 0.77 (0.50, 1.17) | 0.69 (0.43, 1.10) |
| Index cases are confirmed by PCR only | 0.48 (-0.32, 1.29) | 0.02 (-0.33, 0.38) | 0.84 (0.47, 1.49) | 1.13 (0.71, 1.79) | 1.09 (0.65, 1.85) |
| Secondary cases are confirmed by PCR only | **0.78 (0.13, 1.43)** | 0.03 (-0.30, 0.35) | 0.87 (0.51, 1.48) | 0.99 (0.65, 1.50) | 0.88 (0.55, 1.41) |
| Only ancestral strains are circulating in study period | 0.76 (-0.03, 1.55) | 0.07 (-0.31, 0.45) | 1.18 (0.64, 2.17) | 0.76 (0.47, 1.23) | 0.75 (0.43, 1.30) |
| All household contacts were tested | 0.03 (-0.87, 0.93) | 0.28 (-0.04, 0.59) | 1.09 (0.61, 1.94) | 0.85 (0.55, 1.31) | 0.96 (0.58, 1.58) |

\*Estimates based on results from 14 studies

**Table S7.** Pooled estimates for duration of viral shedding for COVID-19 from 11 systematic reviews

| Study | Sampling site | Laboratory method | Pooled estimates (mean/median) | 95% CI | range | I<sup>2</sup> |
| --- | --- | --- | --- | --- | --- | --- |
| Cevik | Upper respiratory tract | PCR | 17.0 | 15.5-18.6 | 6.0- 53.9 |  |
| Cevik | Lower respiratory tract | PCR | 14.6 | 9.3-20.0 | 6.2-22.7 | 97% |
| Cevik | Stool | Viral culture | 17.2 | 14.4-20.1 | 9.8-27.9 | 96.6% |
| Cevik | Serum/blood | Viral culture | 16.6 | 3.6-29.7 | 10.0-23.3 | 99% |
| Fontana | respiratory sources | PCR | 18.4* | 15.5-21.3 | 5.5-53.5 | 98.87% |
| Fontana | Rectal/stool | PCR | 22.1* | 14.4-29.8 | 11-33 | 95.86% |
| Okita | Upper respiratory tract (nasal swab +throat swab) | PCR | 18.29 | 17.00-19.89 | 8.33-39.97 | 99% |
| Okita | sputum | PCR | 23.79 | 20.43-27.16 | 15.50-32.00 | 93% |
| Okita | blood | PCR | 14.60 | 12.16-17.05 | 11.00-17.58 | 88% |
| Okita | stool | PCR | 22.38 | 18.40-26.35 | 10.67-51.40 | 97% |
| Qutub | Respiratory tract | Viral culture | 28.75/11** | 8.5-14.5 |  |  |
| Rahmani | Respiratory tract | PCR | 27.90 | 23.27-32.52 | 7.40-132.00 | 99.1% |
| Xu | respiratory tract (symptomatic cases) | PCR | 11.1±5.8*** |  | 0-24 |  |
| Xu | gastrointestinal tract (symptomatic cases) | PCR | 23.6±8.8*** |  | 10-33 |  |
| Yan | Unrestricted | PCR | 16.8 | 14.8-19.4 |  | 99.56% |
| Yan | stool | PCR | 30.3 | 23.1-39.2 |  | 92.09% |
| Yan | respiratory tract | PCR | 17.5 | 14.9-20.6 |  | 99.67% |
| Yan | Upper respiratory tract | PCR | 17.5 | 14.6-21.0 |  | 99.53% |
| Diaz | stool | PCR | 22* | 19-25 |  |  |
| Chen | (asymptomatic infections) |  | 14.14* | 11.25-17.04 | 11.00-17.25 |  |
| Li | Upper respiratory tract<br>(nasopharyngeal/throat swabs) | PCR | 11.43 | 10.1-12.77 |  | 75.3% |
| Zhang | stool | PCR | 21.8 | 16.4-27.1 |  |  |
| Zhang | respiratory tract | PCR | 14.7 | 9.9-19.5 |  |  |

\*median estimate, \*\*median/grouped median, \*\*\*this study analyzed by cases, and reported mean±SD

**Table S8.** Pooled estimates for duration of viral shedding for COVID-19 in subgroups from 7 systematic reviews

| Study | Sampling site (subgroups) | Laboratory method | Pooled estimates (mean/median) | 95% CI | range | I<sup>2</sup> |
| --- | --- | --- | --- | --- | --- | --- |
| Fontana | respiratory sources (among severely ill patients) | PCR | 19.8* | 16.2-23.5 | 11-31 | 96.42% |
| Fontana | respiratory sources (in mild-to-moderate illness) | PCR | 17.2* | 14.0-20.5 | 8-25 | 95.64% |
| Okita | Upper respiratory tract (nasal swab) | PCR | 19.34 | 16.60-22.07 |  | 99% |
| Okita | Upper respiratory tract (throat swab) | PCR | 17.85 | 16.43-19.26 |  | 98% |
| Okita | Upper respiratory tract (age <60) | PCR | 16.95 | 13.56-20.35 | 8.62-35.67 | 98% |
| Okita | Upper respiratory tract (age ≥60) | PCR | 21.24 | 14.06-28.41 | 8.25-40.33 | 99% |
| Okita | Upper respiratory tract (with comorbidities) | PCR | 20.26 | 17.60-22.92 | 9.67-34.00 | 93% |
| Okita | Upper respiratory tract (without comorbidities) | PCR | 14.66 | 12.63-16.69 | 10.82-27.25 | 85% |
| Okita | Upper respiratory tract (severe patients) | PCR | 20.79 | 18.03-23.55 | 14.00-38.33 | 98% |
| Okita | Upper respiratory tract (nonsevere patients) | PCR | 16.36 | 14.07-18.66 | 8.00-37.33 | 99% |
| Okita | Upper respiratory tract (severe patients) (for studies with mean age ≥40 + comorbidity >30%) | PCR | 21.53 | 17.57-25.50 | 14.00-29.50 | 91% |
| Okita | Upper respiratory tract (severe patients) (for studies with mean age≥40 + comorbidity$>$30%) | PCR | 20.08 | 15.87-24.29 | 13.12-33.67 | 100% |
| Okita | Upper respiratory tract (treated with glucocorticoid) | PCR | 19.72 | 17.92-21.52 | 13.87-33.67 | 92% |
| Okita | Upper respiratory tract (treated without glucocorticoid) | PCR | 15.64 | 14.18-17.10 | 8.33-31.60 | 96% |
| Okita | Upper respiratory tract (treated with glucocorticoid) (for studies with mean age: 30-60 + comorbidity$>$50%) | PCR | 21.98 | 16.48-27.48 | 14.25-33.67 | 94% |
| Okita | Upper respiratory tract (treated without glucocorticoid) (for studies with mean age: 30-60 + comorbidity$>$50%) | PCR | 16.14 | 12.60-19.68 | 13.22-24.44 | 92% |
| Okita | Upper respiratory tract (Asian) | PCR | 18.10 | 16.95-19.25 | 8.33-34.75 | 98% |
| Okita | Upper respiratory tract (European) | PCR | 19.27 | 11.59-26.95 | 8.50-39.97 | 100% |
| Okita | Upper respiratory tract (Asian) (for studies with mean age≥40 + comorbidity$>$40%) | PCR | 20.66 | 18.18-23.14 | 12.00-32.00 | 96% |
| Okita | Upper respiratory tract (European) (for studies with mean age≥40 + comorbidity$>$40%) | PCR | 23.68 | 10.85-36.51 | 13.00-39.97 | 100% |
| Qutub | Respiratory tract (severe patients) | Viral culture | 47.5/20** | 9.0-53.0 |  |  |
| Qutub | Respiratory tract (severe patients were not specified of excluded) | Viral culture | 10/9** | 8.0-13.0 |  |  |
| Qutub | Respiratory tract<br>(immunocompromised patients) | Viral culture | 54.36/20** | 9.0-85.98 |  |  |
| Qutub | Respiratory tract<br>(immunocompromised patients were<br>not specified of excluded) | Viral culture | 11.67/9** | 8.2-13.3 |  |  |
| Rahmani | Noe specific (imunocompetent<br>individuals) | PCR | 26.54 | 21.44-31.64 | 7.40-91.20 | 99.3% |
| Rahmani | Noe specific (immunocompromised<br>individuals) | PCR | 36.28 | 21.93-50.63 | 15.90-132.00 | 94.2% |
| Xu | respiratory tract (asymptomatic cases) | PCR | 9.4±5.1*** |  |  |  |
| Xu | gastrointestinal tract (asymptomatic<br>cases) | PCR | 16.8±9.8*** |  |  |  |
| Yan | Unrestricted (symptomatic cases) | PCR | 19.7 | 17.2-22.7 |  | 99.34% |
| Yan | Unrestricted (asymptomatic cases) | PCR | 10.9 | 8.3-14.3 |  | 98.89% |
| Yan | Unrestricted (severe patients) | PCR | 24.3 | 18.9-31.1 |  | 91.88% |
| Yan | Unrestricted (nonsevere patients) | PCR | 22.8 | 16.4-32.0 |  | 99.81% |
| Yan | Unrestricted (females) | PCR | 19.4 | 9.5-39.4 |  | 93.93% |
| Yan | Unrestricted (males) | PCR | 11.9 | 8.4-16.9 |  | 87.83% |
| Yan | Unrestricted (adults) | PCR | 23.2 | 19.0-28.4 |  | 99.24% |
| Yan | Unrestricted (children) | PCR | 9.9 | 8.1-12.2 |  | 85.74% |
| Yan | Unrestricted (with chronic diseases) | PCR | 24.2 | 19.2-30.2 |  | 84.07% |
| Yan | Unrestricted (without chronic diseases) | PCR | 11.5 | 5.3-25.0 |  | 82.11% |
| Yan | Unrestricted (treated with<br>corticosteroid) | PCR | 28.3 | 25.6-31.2 |  | 0.00% |
| Yan | Unrestricted (treated without corticosteroid) | PCR | 16.2 | 11.5-22.5 |  | 92.27% |
| Yan | Unrestricted (antiviral treatment) | PCR | 17.6 | 13.4-22.2 |  | 98.99% |
| Yan | Unrestricted (mono-antiviral treatment) | PCR | 21.2 | 15.3-29.2 |  | 90.04% |
| Yan | Unrestricted (multiantiviral treatment) | PCR | 20.3 | 13.7-30.3 |  | 99.46% |
| Chen | Unrestricted (asymptomatic infections) | PCR or serum antibody | 14.14* | 11.25-17.04 | 11.00-17.25 |  |

\*median estimate, \*\*median/grouped median, \*\*\*this study analyzed by cases, and reported mean±SD

**Table S9.** Pooled estimates for duration of infectious period for COVID-19 in subgroups from 2 systematic reviews

| Study | Sampling site (subgroups) | Laboratory method | Pooled estimates | 95%CI | range | I<sup>2</sup> |
| --- | --- | --- | --- | --- | --- | --- |
| Rahmani | Replicant competent virus isolation | Viral culture | 7.27 | 5.70-8.84 | 3.40-89.00 | 92.2% |
| Wang | Not specific | Not specific | 6.25 | 5.09-7.51 |  |  |
| Rahmani | Replicant competent virus isolation (imunocompetent individuals) | Viral culture | 6.33 | 4.92-7.75 | 3.00-13.00 | 92.4% |
| Rahmani | Replicant competent virus isolation (immunocompromised individuals) | Viral culture | 29.50 | 12.46-46.53 | 13.80-89.00 | 84.8% |

